# The 501Y.V2 SARS-CoV-2 variant has an intermediate viral load between the 501Y.V1 and the historical variants in nasopharyngeal samples from newly diagnosed COVID-19 patients

**DOI:** 10.1101/2021.03.21.21253498

**Authors:** Elisa Teyssou, Cathia Soulie, Benoit Visseaux, Sidonie Lambert-Niclot, Valentine Ferre, Stéphane Marot, Aude Jary, Sophie Sayon, Karen Zafilaza, Valentin Leducq, Aurélie Schnuriger, Basma Abdi, Marc Wirden, Nadhira Houhou-Fidouh, Charlotte Charpentier, Laurence Morand-Joubert, Sonia Burrel, Diane Descamps, Vincent Calvez, Anne Geneviève Marcelin

## Abstract

The 501Y.V2 and the 501Y.V1 SARS-CoV-2 variants emerged and spread rapidly into the world. We analysed the viral load of 643 nasopharyngeal samples of COVID-19 patients at diagnosis and found that the 501Y.V1 and the 501Y.V2 variants presented a viral load three to ten times and two times higher than the historical variants, respectively.

**Highlights:**

- Viral load in COVID-19 patient nasopharyngeal samples was different between variants.
- The 501Y.V2 variant had a viral load 2 times higher than the historical variants
- The 501Y.V2 variant had a viral load slightly lower than the 501Y.V1
- The 501Y.V1 variant had a viral load 3-10 times higher than the historical variants

---

A rapid spread of the 501Y.V2 (lineage B1.351) into the world and specially in the United Kingdom (UK) has been reported by Wang et al.^1^. As the 501Y.V1 (lineage B1.1.7), which emerged in the UK^2^, this variant notably present specific genetic patterns in the gene encoding the Spike protein (S). It was estimated that the 501Y.V1 presented a greater infectivity (up to 70%) and recent studies suggested a higher viral load (VL) for the 501Y.V1 as compared to the historical SARS-CoV-2^3,4^. Currently, few data are available on the VL of the 501Y.V2. In France, these two variants are actively spreading into the population since late January and currently representing up to 70% of the new positive cases for the 501Y.V1 and 6% for the 501Y.V2 in Paris area.

In this study, we compared the relative VL of the 501Y.V2, with others SARS-CoV-2 variants: the 501Y.V1 and the historical SARS-CoV-2 variants collected from three hospital laboratories in Paris (Pitié-Salpêtrière, Bichat-Claude Bernard and Saint-Antoine/Trousseau hospitals). A total of 643 RT-PCR SARS-CoV-2 positive nasopharyngeal samples collected at diagnosis were screened to assess SARS-CoV- 2 viral lineages with the TaqPath™ COVID-19 RT-PCR (ThermoFisher, Waltham, USA) and the VirSNiP SARS-CoV-2 Spike E484K (TIB Molbiol, Berlin, Germany). The TaqPath COVID-19 test amplified three target genes of the virus (ORF1ab, N, and S). The 501Y.V1 presents a specific deletion in the S gene, *69-70del*, which results in a failure of its detection by this assay. Then, a Sanger sequencing amplifying the RBD region was realised to distinct between the 501Y.V2 and a new variant, B1.1.248, (N501Y and E484K mutations). The relative VL (copies/ml) was assessed by linear regression with a standard curve established from a SARS-CoV-2 positive nasopharyngeal sample quantified by Droplet-Digital^tm^ PCR (Bio-Rad). An ANOVA with a multiple comparison test was performed with the STATVIEW software.

We analysed the results from 643 SARS-CoV-2 infected patients: 332 historical SARS-CoV-2, 249 501Y.V1 and 62 501Y.V2 which presented similar median age at 57 years [38-76] and sex ratio with 53% of female. For the N gene, the 501Y.V2 presented a relative VL two times higher (median 2.32e+7 copies/ml [8.52e+5-2.40e+8]) than the historical variants (median 1.05e+7 [1.81e+5-1.41e+8]) (p<0.0001). Moreover, the 501Y.V1 (median 1.12e+8 [1.34e+6-1.19e+9]) presented a relative VL ten times higher than the historical (p<0.0001) (Fig.1A). For the ORF1ab gene, the 501Y.V2 presented also a relative VL (median: 2.69e+7 [6.47e+5-2.40e+8]) two times higher than the historical variants (median: 1.18e+7 [1.77e+5-1.60e+8]) (p<0.005). Unlike the N gene, no statistical difference was found between the 501Y.V2 and the 501Y.V1 (median: 3.80e+7 [6.14e+5-4.85e+8]) which also presented a relative VL two times higher than the historical variants (p<0.0001) (Fig.1B).

**Figure 1:**
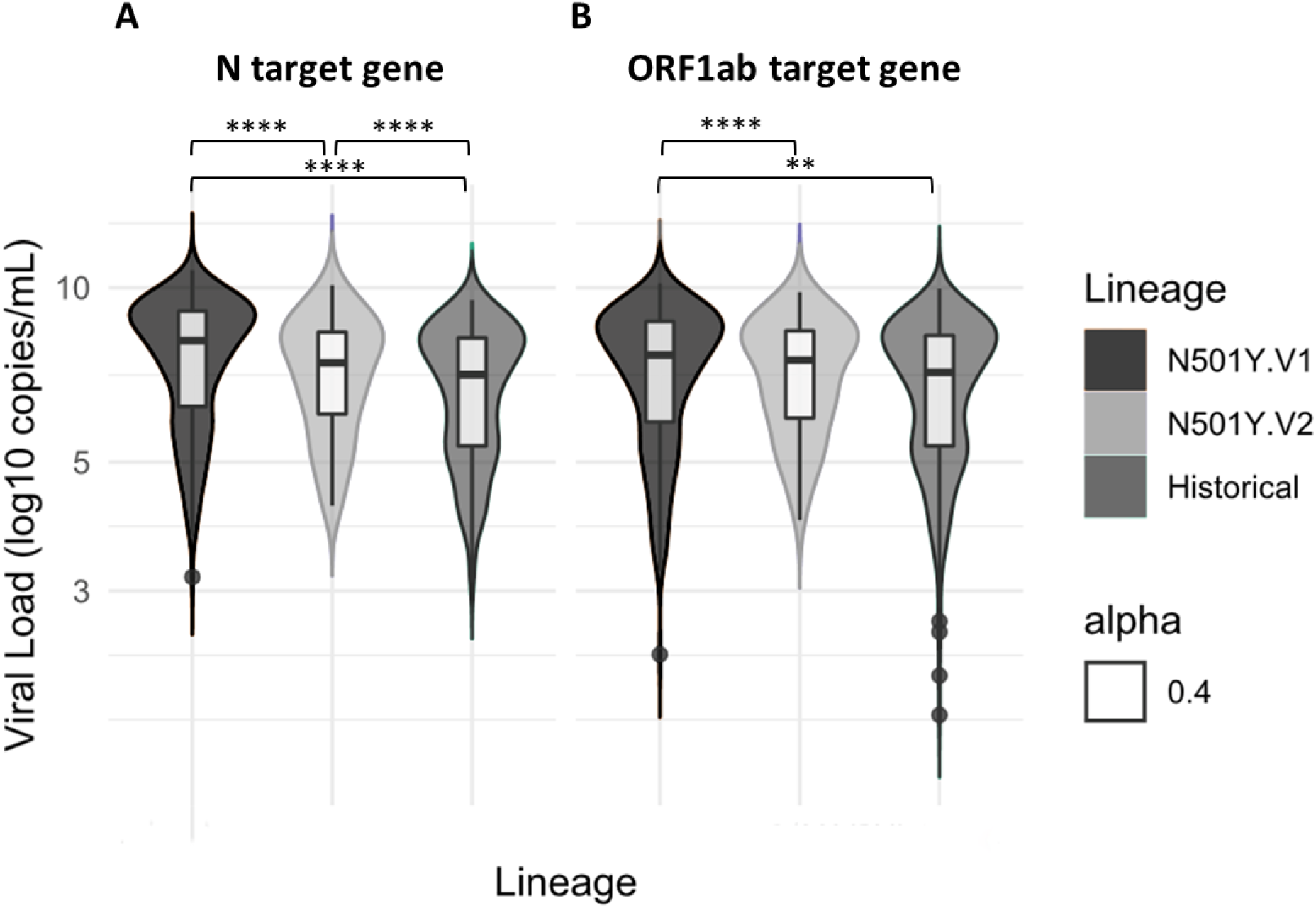
501Y.V2 and 501Y.V1 are associated with a higher viral load than historical SARS-CoV-2.

The graph presents the median and the minimal to maximal of the relative VL in log copies/ml of the three groups for the N gene (A) and for the ORF1ab gene (B). The median of the relative VL was higher for the 501Y.V2 and the 501Y.V1 than the historical SARS-CoV-2. **p<0.005; ****p<0.0001.

Our results showed significant differences of VL between these three SARS-CoV-2 variants. Indeed, we found that both new 501Y.V1 and 501Y.V2 variants have statistically higher nasopharyngeal relative VL at diagnosis than the historical lineages.

In other respiratory diseases like SARS and Flue, it has been shown that the level of VL influences outcomes of the disease. In Flue, a higher VL was observed for H5N1- infected patients compared to H3N2 and H1N1-infected patients and was associated with a strongest severity of the disease^5^. In the SARS-CoV a high VL was associated with the severity of symptoms and was shown to be a good indicator of the respiratory failure and death^6^.

A similar pattern was observed with the SARS-CoV-2 virus. A higher VL at the first SARS-CoV-2 RT-PCR testing could be associated with a longer viral persistence of the virus, the contagious condition of patient and can be used as a predictive indicator of the severity of the disease^4,7^. Further studies are required to confirm potential higher infectiousness and higher severity of the disease caused by these two emerging variants.

However, our study reinforces the hypothesis that the 501Y.V1 variant is more infectious than the historical SARS-CoV-2^3,4^ and thorough this hypothesis for the 501Y.V2 which begins to spread in France.

Our study is limited by the fact that we were focused on the relative VL at one time, which could be influenced by the variation of the nasopharyngeal swab technique or by the timeline of the epidemic. With the aim to limit this fluctuation, we only collected sample for the first presentation test and with the same SARS-CoV-2 assay at the same period (December 2020 to February 2021). Another limitation is the small number of positive cases for the 501Y.V2 compared to the two other strains principally due to the only few positive cases in Paris area at that time. However, given the co-circulation of these two variants in France and their high transmissibility, it is important to strengthen the SARS-CoV-2 genomic surveillance to follow the evolution of their respective prevalence.

The two new variants seem to present a higher VL compared to the historical SARS-CoV-2 which might arise from the fact that they share the same N501Y mutation probably leading to a better infectivity^8,9^. However, the 501Y.V2 seems to be associated with a lower VL compared to the 501Y.V1 for the N gene, a difference which fades for the ORF1ab gene. This could be due to their other specific mutations. Considering the 501Y.V2, it does not seem, contrary to the 501Y.V1, that the Spike RBD has higher affinity for ACE2 as compared to the reference Spike. If indeed the transmission is enhanced, this could be due to mutation outside the direct ACE2–Spike interface^10^. Moreover, it has been shown that an active viral replication was associated with the transcription of subgenomic viral RNA and that this active replication occurs during the first days after the onset of symptoms^11^. As we collected only the first presentation test sample, which suggest the presence of an active viral replication, the difference between the N and the ORF1ab genes could be explained by the presence of these subgenomic viral RNA.

Our study brings evidence of increased infectivity for the 501Y.V1 and new proofs for the little-known 501Y.V2 variant infectivity that could explain their propagation velocity worldwide.

## Data Availability

all the data in this manuscript are available. The data are presented on an excel format.

## Acknowledgements

The authors acknowledged all the member of the Pitié-Salpêtrière, Bichat and Saint-Antoine/Trousseau Virology departments for their collaboration and clinicians of the three hospitals for their implication in the SARS-CoV-2 patient care. We thank the ANRS-MIE (Agence Nationale de Recherches sur le SIDA et les hépatites virales- Maladies Infectieuses Emergentes) (AC43, Medical Virology) for its support.

